# Comparative cortical transcriptomic profiling of Alzheimer’s disease, vascular dementia and mixed dementia

**DOI:** 10.1101/2025.09.11.25335564

**Authors:** Frances T. W. Lim, Yuek Ling Chai, Jasinda H. Lee, Clara Y. B. Low, Paul T. Francis, Clive Ballard, Raj N. Kalaria, Brian K. Kennedy, Christopher P. Chen, Tau Ming Liew, Mitchell K. P. Lai, Michelle G. K. Tan

## Abstract

Alzheimer’s disease (AD) and vascular dementia (VaD) are two of the commonest causes of dementia worldwide. While AD is characterized by amyloid plaque and neurofibrillary tangle formation, and VaD is characterized by cerebrovascular disease (CeVD), these pathophysiological processes frequently coexist, leading to mixed AD+VaD dementia (MIX). At present, it is unclear which of the multiple gene expression changes observed in MIX brains are driven by AD versus VaD processes. In this study, postmortem neocortical tissues of AD (n=9), VaD (n=9), and MIX (n=10), together with age-matched controls (CTRL, n=10), underwent transcriptome profiling in conjunction with pathway analyses using an established exon-microarray platform. Transcriptome profiling showed that up- and down-regulated genes are mainly associated with vascular dysfunction and neurodegeneration, respectively. VaD manifested the least number of differentially expressed genes (DEG) amongst the diagnostic groups, showing similar but relatively less pronounced changes in common dementia-associated pathways. MIX shared high similarities with AD in gene expression profiles and dysregulated canonical pathways, with additional, MIX-specific DEG and dysregulated pathways suggestive of additive or emergent deleterious effects from AD and cerebrovascular pathologies. Our study provided a genome-wide overview of the gene expression landscape for AD, VaD and MIX, enabling the identification of common and disease-specific pathophysiological processes which inform further studies into the complex interactions between AD and CeVD.

## 1 BACKGROUND

Dementia is a major unmet healthcare need in aging populations worldwide [1]. The leading cause of dementia in older persons is Alzheimer’s disease (AD), characterized neuropathologically by aggregated β-amyloid-containing amyloid plaques (AP) and neurofibrillary tangles (NFT) formed from paired helical filaments of hyper-phoshorylated tau [2]. Vascular dementia (VaD), the second leading cause of age-related dementia, presents with cerebrovascular diseases (CeVD) resulting in cortical hypoperfusion, ischemia, cerebral microbleeds, white matter lesions, and infarction (cortical, lacunar and microinfarcts) [3, 4]. Of the wide range of CeVDs, small vessel diseases underlie the commonest, subcortical ischemic form of VaD which predominantly involve the basal ganglia, cerebral white matter and the brainstem, resulting in a clinical presentation of dementia and significant cognitive deficits including executive dysfunction [5, 6].

Despite having apparently distinct pathogenic mechanisms, AD and VaD share several risk factors, including older age, hypertension, hyperlipidemia and diabetes [7, 8]. Furthermore, both AD and VaD processes contribute to white matter lesions, microinfarcts, and brain atrophy, eventually leading to cognitive decline and dementia [8]. Indeed, the frequent co-existence of AD and VaD pathologies of sufficient severity to independently underlie clinical dementia gives rise to mixed AD+VaD dementia (MIX) [9–11]. Even in people diagnosed only with AD, small vessel CeVDs are often concomitant to amyloid pathology, and are known to exert additive or synergistic deleterious effects [8, 12, 13]. However, it remains unclear to what extent AD-associated pathology and CeVD contribute to the transcriptomic landscape of MIX.

While genome-wide association studies and biofluid-based “-omics” studies have compared AD to CeVD-driven alterations [14–17], there is limited research on differentially expressed genes (DEG) in AD postmortem cortex [18, 19], and even fewer on VaD. Only one major study on global transcriptomic analysis between AD and VaD has reported that AD processes drove significantly more DEGs (76%) than VaD (28%) [20, 21], while direct comparisons among AD, VaD and MIX are not yet available. Hence, in this study, we used a high-throughput, established transcriptomic profiling platform to generate a comprehensive gene expression landscape using neocortical tissue derived from well-characterized cohort of AD, VaD, MIX and older, cognitively unimpaired control participants (CTRL) to identify key biological processes associated with each dementia subtype.

## 2 METHODS

### 2.1 Subjects, clinical and pathologic assessments

Postmortem tissues predominantly from the temporal cortex (Brodmann area BA22) were obtained from the Thomas Willis Oxford Brain Collection, the Newcastle Brain Tissue Resource, and the London Neurodegenerative Diseases Brain Bank, all of which are part of the Brains for Dementia Research initiative (https://bdr.alzheimersresearchuk.org/) jointly supported by Alzheimer’s Society and Alzheimer’s Research UK. The collection and investigations of brain tissues have received Institutional Review Board approval in both the UK (08/H1010/4) and Singapore (NUS 12-062E) institutions, and informed consent for the removal of brain was obtained from the participants’ next-of-kin. Before death, all participants were recruited into longitudinal studies of dementia, which included annual cognitive assessments using the Mini-Mental State Examination (MMSE)[22] as an indicator of severity of cognitive impairment, with clinical diagnosis of dementia further based on the Diagnostic and Statistical Manual of Mental Disorders (Fourth Edition) criteria. AD neuropathological assessments included the National Institute on Aging-Alzheimer’s Association guidelines with phases of Aβ deposition, Braak stages and Consortium to Establish a Registry for Alzheimer’s disease (CERAD) scores [23–25]. Diagnosis of vascular dementia was based on NINDS-AIREN operationalized criteria [26]. Neuropathological confirmation of subcortical ischemic VaD was based on findings of small vessel diseases in subcortical structures, including microinfarcts, lacunes and white matter lesions [27]. Participants with cortical infarcts were not selected for this study to avoid gene expression measurements on infarcted neocortical tissue. Frozen brain chunks (≈1cm^3^) contralateral to the hemisphere used for neuropathologic studies was thawed on ice, dissected free of meninges and white matter, and processed independently for RNA isolation (see below), as well as homogenized in Tris-HCL buffer for enzyme-linked immunosorbent assays (ELISA) of tau phosphorylated at serine-396 (pS396 Tau) and total Tau as previously reported [28, 29] to derive values for ratios of pS396 Tau to total Tau.

### 2.2 Transcriptome profiling using Affymetrix Gene arrays

Total RNA derived from the neocortex of n = 10 CTRL, 9 AD, 9 VaD and 10 MIX (mean ± SD RNA Integrity Number = 5.06 ± 0.89 as determined by an Agilent Bioanalyzer, Agilent Technologies, Santa Clara, CA, USA) were processed for high-throughput transcriptome profiling. Briefly, 50 ng total RNA was reverse transcribed and amplified using Applause™ WT-Amp ST kit (NuGEN Technologies, Rosemount, MN, USA), followed by fragmentation and biotin labelling using Encore® Biotin module (NuGEN) and hybridized to Affymetrix GeneChip® Human Gene 1.0 ST (ThermoFisher Scientific, Waltham, MA, USA) at 45°C for 16 hours with rotation at 60 rpm, then washed and stained in a Affymetrix Fluidics Station 450. Gene chips were scanned by an Affymetrix 3000 7G scanner. CEL files were generated from Affymetrix GCOS Software and imported into Partek Genomics Suite® 7.0 (http://www.partek.com/partekgs) software (Illumina, San Diego, CA, USA) for analysis. Probe summarization and probe-set normalization were performed using Robust Multi-Chip Average (RMA) which included RMA background corrections, quantile normalization, log-2 base transformation and summarization by Median Polish. Exon-level analysis under exon expression workflow was performed with default parameters. Differentially expressed probesets were processed by alternative Splice ANOVA, using gender as covariate to determine differentially expressed transcripts in AD, VaD and MIX versus CTRL, based on False Discovery Rate (FDR) q < 0.1 and fold change (FC) cutoff = 1.5. After removing nil annotation or repeated annotation data, the full list of differentially expressed genes (DEGs) presented as Transcript Cluster ID for downstream analysis was obtained (see **Supplementary Table S1**). To facilitate principal component analysis (PCA) and hierarchical clustering analysis, gene-level analyses were performed to summarize exons to genes using Partek’s software. Subsequently, up- and down-regulated DEGs were analyzed for enrichment of gene ontology (GO) and Kyto Encyclopedia of Genes and Genomes (KEGG) pathway database using DAVID Bioinformatics Resources (https://davidbioinformatics.nih.gov/) with Benjamini’s enrichment p-values adjustment for multiple testing using FDR of 5%.

### 2.3 Functional analysis using Ingenuity pathway analyses (IPA)

Functional implications of DEGs were analyzed by Qiagen’s Ingenuity® Pathway analyses (IPA, https://digitalinsights.qiagen.com/) to identify significant inhibition or activation of canonical pathways, as well as to detect involvement of physiological or pathophysiological processes in dementia. Firstly, up- or down-regulated DEGs in AD, VaD and MIX were analyzed independently by IPA core analysis, followed by comparison analysis.

For core analysis, p-values were calculated using right-tailed Fisher’s Exact Tests to assess the likelihood that the association between sets of biomolecules derived from DEG and specific processes, pathways, or transcription neighborhoods is due to chance. For comparison analysis, p-value adjusted by Benjamini-Hochberg’s FDR procedure (B-H p-value) was used to measure the significance of each entity in association with the dataset, where - log (B-H p-value) >1.3 corresponds to a B-H p-value <0.05. Z-scores of matches between expected relationship direction and observed gene changes were calculated for each entity based on the number of known target genes of interest, pathway or function, expression changes of these target genes, as well as concordance with published literature. Z-scores >2 or < -2 were considered significant for states of activation and inhibition, respectively.

### 2.4. Reverse transcription quantitative polymerase chain reaction (RT-qPCR)

Validation of gene expression were carried out for selected DEGs in each dementia group by RT-qPCR. Briefly, 2μg of RNA was reverse-transcribed to cDNA using High-Capacity cDNA RT kit (Applied Biosystems, Foster City, CA, USA) in accordance with manufacturer’s protocol. Semi-quantitative measurements of gene expression were performed on CFX96^TM^ Real-time PCR system (Biorad Laboratories, Hercules, CA, USA) using GoTaq® qPCR Master Mix (Promega, Madison, WI, USA). The primer sequences and PCR conditions used in this study are listed in **Supplementary Table S2**. All RT-qPCR assays were performed in duplicate. Standard curves of each gene were generated independently by 10x serial dilution of template DNA. The relative signal intensity of each sample was calculated according to the corresponding standard curve. Normalization was performed in each sample by dividing the relative signal intensity of gene of interest to geometric mean of β-actin, GAPDH and 18S rRNA.

### 2.5 Statistical analyses

Statistical analyses were performed using the SPSS Statistics software (version 29, International Business Machines Corporation, Armonk, NY, USA), with additional graphing performed on PRISM software (Version 6, GraphPad, San Diego, CA, USA). Group differences in output values analyzed by one-way analysis of variance (ANOVA) with Bonferroni’s *post-hoc* tests, or Kruskal-Wallis test with *post-hoc* Dunn’s tests when measurement variables did not meet normality assumptions. Determination of significant DEGs was based on FDR q < 0.1 using the Partek software (see **Section 2.2**), while DAVID and IPA analyses were based on B-H adjusted p < 0.05 (see **Section 2.3**). For other analyses, two-tailed p-values of < 0.05 were considered statistically significant.

## 3 RESULTS

### 3.1 Demographic and disease variables of study participants

Demographic and disease parameters of the study participants are listed in **Table 1**. Non-dementia control, AD, VaD and MIX participants did not differ in age, gender distribution, and postmortem interval (ANOVA F test *p* > 0.05). For participants classified as VaD, none had Braak stage > II tangle burden or more than “sparse” plaque counts by CERAD criteria. MIX participants were classified as VaD as well as having “probable” or “definite” AD diagnosis by CERAD criteria, with Braak staging for both MIX and AD participants were all > III/IV. In corroboration with neuropathological assessments, pS396Tau : Total tau as a neurochemical marker of neurofibrillary tangle burden [28, 29] were higher in both AD and MIX when compared with aged CTRL and VaD (**Table 1**). Furthermore, AD and MIX manifested more severe dementia as shown by the lowest pre-death MMSE scores, with VaD (predominantly of the subcortical ischemic type) showing higher MMSE than MIX and AD, but lower than CTRL, suggesting relatively mild cognitive impairment in the VaD subgroup (**Table 1**), consistent with previous findings [6].

**Table 1.** Demographics and disease variables in the study cohort.

| **Variables** | **CTRL**<br>**(n=10)** | **AD**<br>**(n=9)** | **VaD**<br>**(n=9)** | **MIX**<br>**(n=10)** |
| --- | --- | --- | --- | --- |
| *Gender (male/female)* | 6M/ 4F | 6M/ 3F | 6M/ 3F | 6M/ 4F |
| *Age at death (yr)* | 83.0±2.4 | 71.3±2.9 | 80.8±3.5 | 77.6±3.3 |
| *Postmortem interval (h)* | 29.9±5.4 | 62.9±9.9 | 44.7±10.4 | 41.1±7.8 |
| *MMSE before death* | 28.5±0.3 | 5.5±2.6* | 23.2±2.0^ | 11.7±2.5* |
| *pS396Tau : total Tau* | 0.018±0.004 | 1.998±0.348* | 0.008±0.003^# | 0.397±0.180* |

Data are presented as mean± S.E.M. AD, Alzheimer's disease; CTRL, control; MIX, mixed AD and VaD; MMSE, Mini-Mental State Examination; VaD, vascular dementia. pS396Tau : total Tau was measured in the temporal cortex (BA22). Significant differences as compared to \*CTRL, ^AD or #MIX were determined by Kruskal-Wallis ANOVA with *post-hoc* Dunn's test, $p<0.05$ .

### 3.2 Gene expression profiles of AD, VaD and MIX

DEGs for each dementia group (AD, VaD and MIX) were derived by comparisons with CTRL (see **Supplementary Table S1**). **Figure 1A** depicts proportional Venn diagrams of up-and down-regulated DEGs between AD, VaD and MIX. 18.9% up-regulated (162/857) and 34.7% down-regulated (501/1445) DEGs were found in common in all dementia groups (white region), while VaD had the lowest number of DEGs among the groups (orange region). A higher degree of overlap was found between AD and MIX (cyan region), than between VaD and MIX (yellow region). Likewise, a lower level of similarity was also detected between AD and VaD (pink region). **Figure 1B** showed DEGs considered significant in each dementia group based on fold change cut-off of 1.5 and FDR q < 0.1. Using PCA, DEGs were confirmed to be distributed into four distinctive clusters corresponding to the diagnostic groups (**Figure 1C**). CTRLS (red) showed tight clustering and were well-segregated from the dementia groups, while AD (blue) and VaD (amber) seemed to have wider sample variations as shown by larger ellipsoid coverage areas. Although each of the three dementia groups could be clustered with minimal overlap, they remained close to one another on the PCA space, suggesting a degree of similarity in gene alteration patterns. Next, we performed unsupervised hierarchical clustering analysis for the top 100 DEGs within the dementia groups (**Figure 1D**). CTRL samples showed consistent gene expression pattern within the group and clustered with some VaD samples within the upper branch of vertical dendrogram. The lower branch, namely dementia cluster, contained sub-branches for VaD, AD and MIX, as well as a mixture of MIX and AD, suggesting that MIX and AD shared a high degree of gene alterations, whereas VaD showed relatively lower gene alterations compared with AD and MIX.

**Figure 1.**
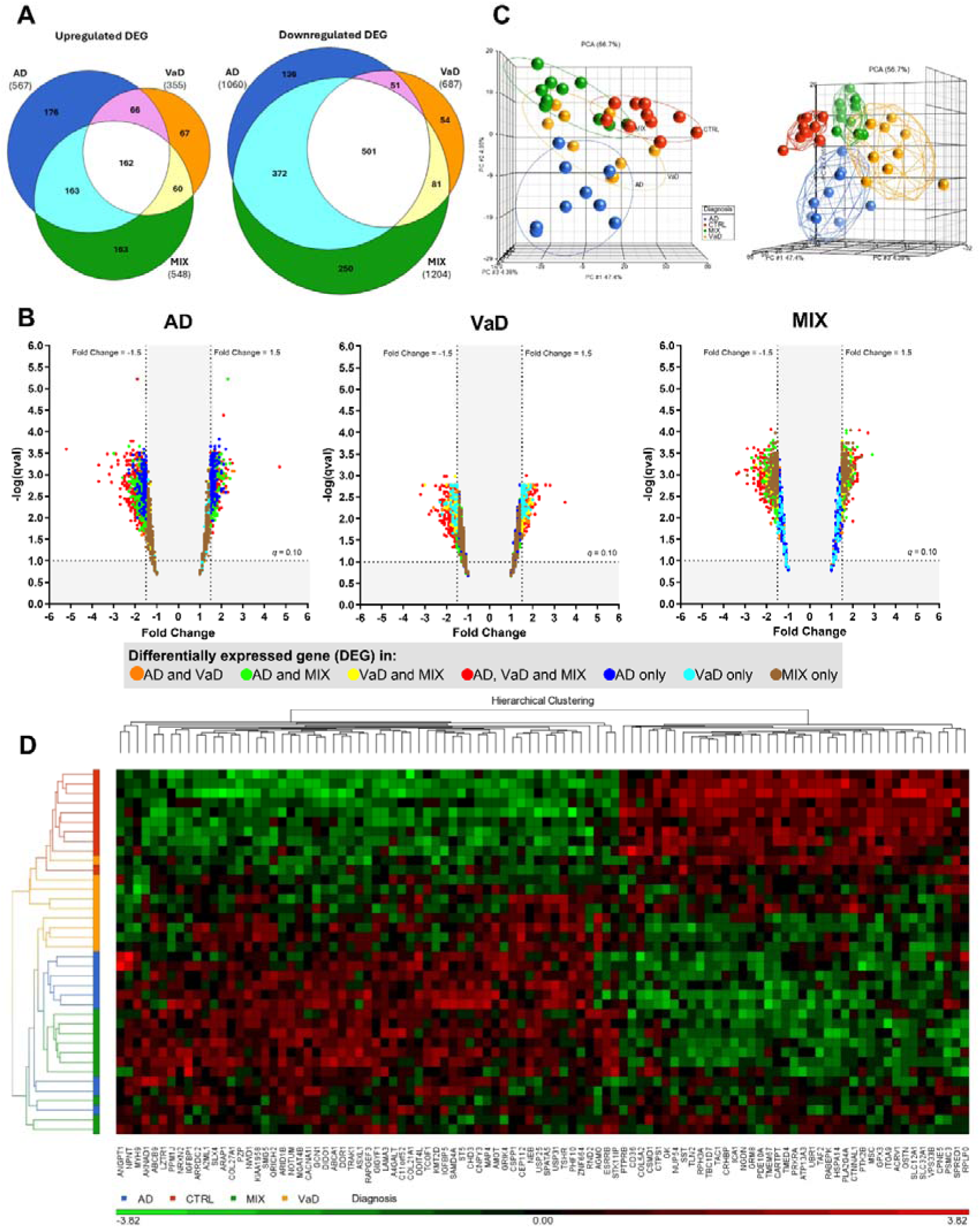
Distribution, principal component analysis and heatmap of differentially expressed genes in AD, VaD and MIX. (**A**) Proportional Venn diagrams summarized similarities and differences of gene expression changes between Alzheimer’s disease (AD), vascular dementia (VaD) as well as mixed (AD and VaD) dementia (MIX). Numbers of up- and downregulated differentially expressed genes (DEGs) were displayed in the respective regions of the three-circle scaled Venn diagrams. The complete list of DEGs can be found in **Supplementary Table S1**. (**B**) Volcano plots of DEGs in each dementia group set at cut-off fold change > 1.5 and FDR q < 0.10. Each DEG is color-coded based on detection in different combinations of dementia groups. (**C**) 3-dimension principal component analysis (PCA) plots demonstrate the segregation of Control (CTRL), AD, VaD and MIX groups based on DEGs. Each object represents a subject color-coded for respective diagnostic groups (red for CTRL, blue for AD, amber for VaD and green for MIX). The same PCA plot was depicted from two different angles to provide improved visualization of group segregation. Ellipses on the left figure and ellipsoids on the right figure were drawn for each group to highlight the segregation degree. (**D**) Unsupervised hierarchical clustering heatmap displaying overall expression patterns of the top 100 DEGs among diagnostic groups (CTRL n=10, AD n=9, VaD n=9 and MIX n=10) analyzed using Euclidean minimum distance clustering algorithm to produce the heatmap with dendrograms for samples (left panel) and genes (top panel). The gene expression levels are shown with the rows (samples) and column (genes) in cluster order and scaled so that mean expression is zero and standard deviation is one. Samples with higher expression of a given gene were marked in red and those with lower expression were marked in green. Corresponding gene names are listed at bottom of each column.

### 3.3 RT-qPCR validation of selected DEGs

In view of the presence of both shared and unique DEGs among AD, VaD and MIX, we selected nine transcriptomics-derived DEGs for validation with RT-qPCR, of which five (*ITPKB*, *GFAP*, *VCAN*, *NPNT*, *HIF3A*) showed significant correlation, one (*RFTN2*) showed marginal correlation, while three (*SLC9A2*, *MRC1*, *DPP4*) did not correlate with corresponding RT-qPCR derived expression (see **Supplementary Figure S1**). Several of the positively correlated genes are involved in well-established pathophysiological processes. For example, astrogliosis, as indicated by increased GFAP expression, has been recognized to be strongly associated with AD pathology [30–32]. In our study, increased *GFAP* was only significant in AD and MIX, but not in VaD as shown by both gene array (**Supplementary Table S1**) and RT-qPCR data (**Figure 2A**). In contrast, *ITPKB* (inositol triphosphate 3-kinase B) is a potential signaling modulator known to exacerbate AD-related pathology, and has been reported to be upregulated in both postmortem AD and AD mouse model brains [33, 34]. In our study, while *ITPKB* was found to be significantly upregulated in AD and MIX, but not VaD with gene array (**Supplementary Table S1**), RT-qPCR confirmed significant increase of *ITPKB* expression in AD when compared with CTRL and VaD, but increases in MIX did not reach statistical significance (**Figure 2B**).

**Figure 2.**
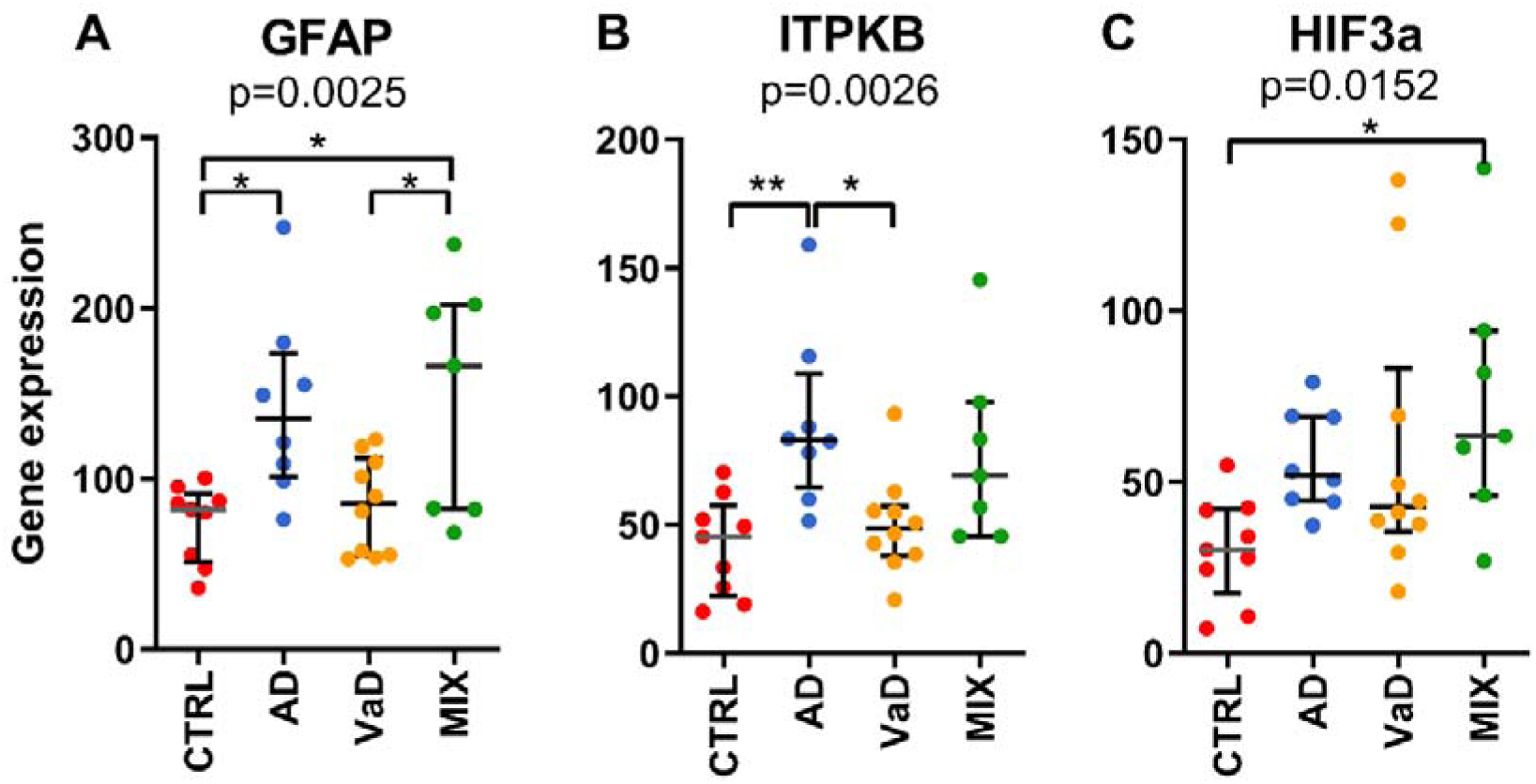
RT-qPCR validation of selected differentially expressed genes in AD, VaD and MIX. Genes of interest were selected from the list of DEGs for validation by qPCR. Dot plots of CTRL (n=9, red), AD (n=8, blue), VaD (n=10, amber) and MIX (n=7, green) showed Log_2_-transformed relative expression of (**A**) *GFAP* (glial fibrillary acidic protein gene); (**B**) *ITPKB* (Inositol-trisphosphate 3-kinase B gene) and (**C**) *HIF3A* (hypoxia-inducible factor 3 alpha gene), with Kruskal-Wallis ANOVA p values listed below each gene name, and horizontal lines in each group indicating median and interquartile range. *p<0.05, **p<0.01, significant difference between groups using Dunn’s *post-hoc* tests.

Among upregulated DEGs in VaD and MIX, we were particularly interested in hypoxia inducible factor 3 alpha subunit (*HIF3A)*, as its expression is known to be increased under hypoxic conditions in endothelial and vascular smooth muscle cells [35], where it acts as a negative regulator for the vascular endothelial (VE)-cadherin [36]. We found that MIX had significant upregulation of *HIF3A*, whereas trends towards increases were detected in VaD and AD (**Figure 2C**), suggesting modulation of *HIF3A* could be associated with the hypoperfusion detected in dementia.

### 3.4 Pathway enrichment analyses of up- and down-regulated DEGs

We observed that both up- and down-regulated DEGs associated with enriched GO and KEGG pathways with distinct functional links (**Supplementary Table S3**). Upregulated DEGs were mainly associated with extracellular matrix organization, microtubule-based movement, positive regulation of GTPase activity and ECM-receptor interactions, likely contributing or responding to neurovascular injury and dysfunction. On the other hand, downregulated DEG were highly associated with synaptic transmission, ion transmembrane transport, neurotransmitter secretion, mitochondrion, and involved in synaptic vesicle cycles, oxidative phosphorylation and metabolic pathways, likely indicating neurodegenerative processes.

We implemented two strategies for IPA pathway core analyses to study the impacts of DEG on potential pathological consequences. First, separate analyses of up- and down-regulated DEGs were performed, with the respective top 10 most significantly altered canonical pathways in AD, VaD and MIX listed in **Figure 3**. Among the top 10 upregulated DEG-driven pathways, caveolar-mediated endocytosis signaling was the only one in common among the dementia groups (see **Figure 3** left panel). Half of the altered pathways in AD and MIX overlapped, including integrin signaling, cardiac hypertrophy signaling/(enhanced) and signaling by Rho family GTPases, likely associated with the dysfunction of the neurovascular unit (NVU) [37–42]. Besides inhibition of PTEN signaling, distinct altered pathways found in VaD were associated with hepatic fibrosis and the GP6 signaling pathway involving various extracellular matrix (ECM) genes like collagens and laminins (**Supplementary Table S4**), possibly associated with dysregulation of vascular endothelium [39, 43, 44].

**Figure 3.**
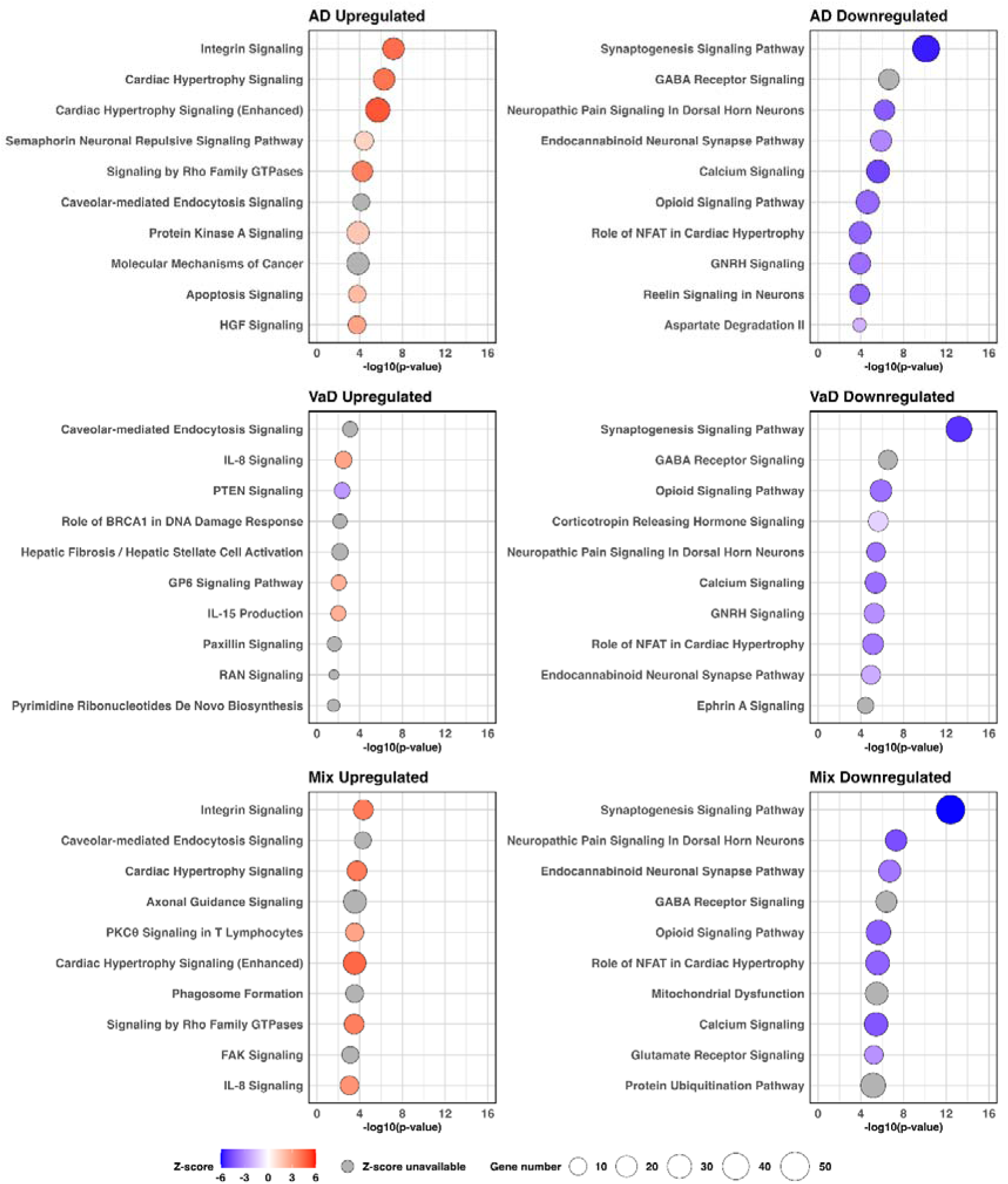
Top 10 altered canonical pathways in AD, VaD and MIX based on up- or downregulated DEGs. Functional implications of up- and down-regulated DEG in AD, VaD and MIX were analyzed using the Qiagen Ingenuity Pathway Analysis (IPA) knowledge base to identify significantly altered canonical pathways. Top 10 significantly altered canonical pathways from each group were displayed as ball charts, with colour indicating Z-score, size indicating number of detected DEGs, and x-axis showing -log_10_(p-value). The complete list of altered canonical pathways in each group and corresponding DEGs can be found in **Supplementary Table S4**.

On the other hand, downregulated DEG-driven pathways appeared to be more conserved, with seven of ten found in all dementia groups (**Figure 3** right panel). These pathways were mainly associated with dysregulation of neuronal functions including synaptogenesis, GABA receptor signaling, neuropathic pain signaling, endocannabinoid neuronal synapse pathway, calcium signaling and opioid signaling pathway, consistent with the neurodegenerative processes.

### 3.5 Significantly altered canonical pathways in MIX demonstrated higher degree of similarity with AD than with VaD

The second strategy of pathway enrichment analysis included all DEGs for AD, VaD and MIX, followed by determination of significant differences of pathway alterations amongst the groups using BH-adjusted p-values to correct for false discovery. With this approach, 59, 25 and 82 significantly altered canonical pathways were identified in AD, VaD and MIX, respectively. The similarities and differences of altered pathways in each group were summarized by proportional Venn diagrams (**Figure 4A**). The top 20 altered canonical pathways were displayed by -log_10_(BH p-value) heat maps sorted by the sum of significance scores, alongside the corresponding Activation Z-score heat maps, showing predicted stages of activation or inhibition (**Figure 4B**). In summary, 18% (17/97) of altered pathways were found in all three dementia diagnostic groups, with more than half of the pathways associated with neurological deficits and overlapping with altered pathways of downregulated DEGs (**Figure 4B** and **Supplementary Table S5**), suggesting that neurodegeneration is a common characteristic of AD, VaD and MIX irrespective of underlying pathologies. Furthermore, neuroinflammation is also a well-established feature of all there dementia groups [45–47], and is corroborated by our finding of neuroinflammatory signaling as another commonly altered pathway (**Supplementary Table S5**). Other pathways found to be altered in all three dementia groups included GP6 signaling, Ephrin A signaling and signaling by Rho family GTPases, reported to be directly or indirectly associated with vascular function or modulation of the neurovascular unit [40, 48, 49].

**Figure 4.**
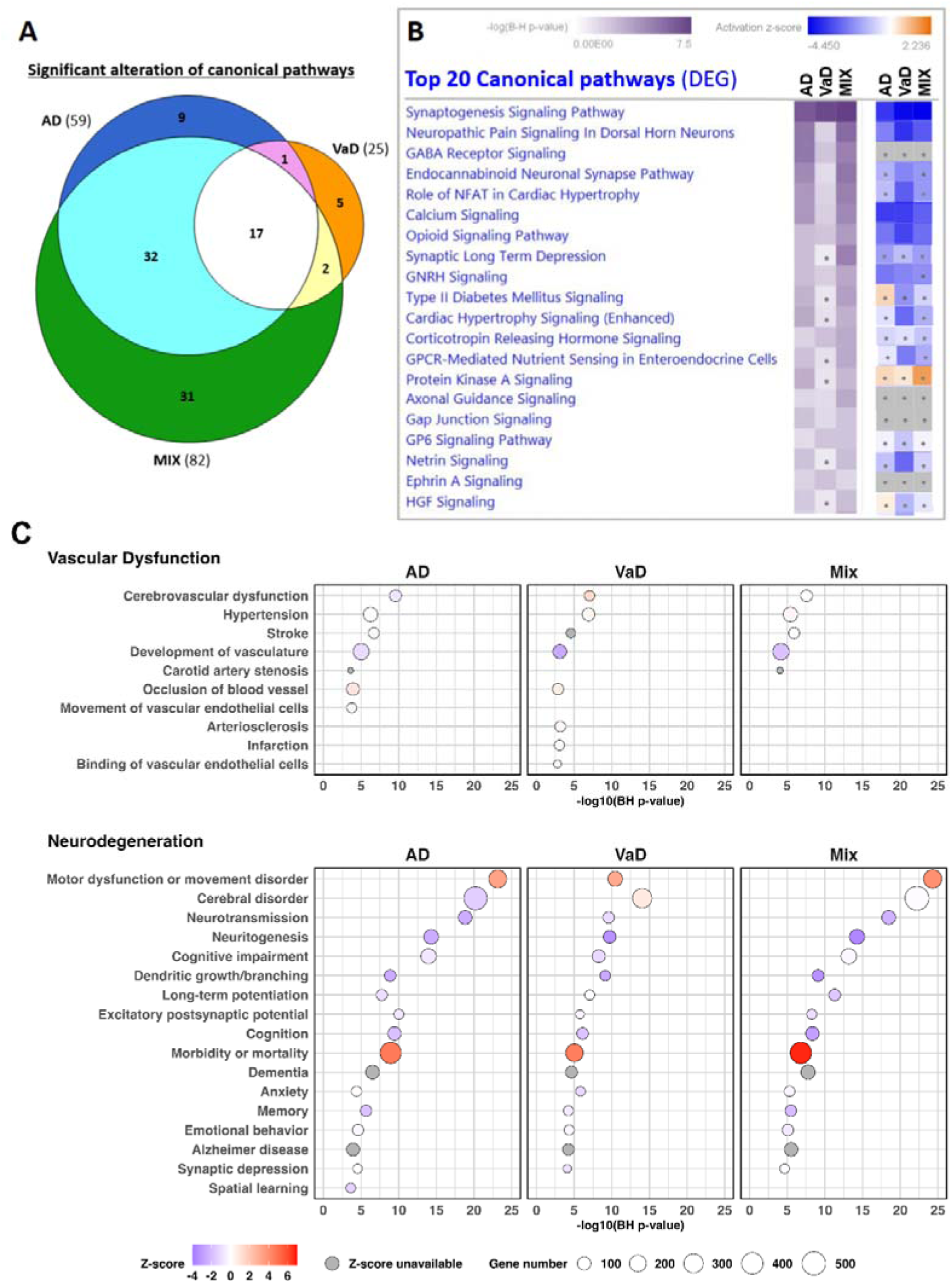
Comparisons of significantly altered canonical pathways in AD, VaD and MIX. Significantly altered canonical pathways in AD, VaD and MIX identified using IPA with -log (B-H p-value) >1.3. (**A**) Proportional Venn diagrams summarizing the numbers of significantly altered canonical pathways with respect to the overlaps and disimilarities among the groups. (**B**) Top 20 altered canonical pathways sorted by the sum of -log (B-H p-value) scores were listed by heat maps using purple color gradient. Activation *Z*-score heat maps are shown side-by-side to infer likely activation states (positive value, indicated by orange color gradient), inhibition states (negative value, indicated by blue color gradient) or unpredictable (grey, NaN). Dots in heat map represent entities of -log (B-H p-value) <1.3 or *Z*-score < |2|. The complete list of altered canonical pathways and corresponding DEGs can be found in **Supplementary Table S5**. (**C**) IPA detection of biofunctions and diseases associated with DEGs displayed as ball charts, with colour indicating Z-score, size indicating number of detected DEGs, and x-axis showing -log_10_(B-H p-value). The complete list of detected diseases and biofunctions as well as corresponding DEGs can be found in **Supplementary Table S6**.

Consistently, we observed a substantial overlap of dysregulated pathways between AD and MIX (33%, 32/97, cyan region in **Figure 4A**) which included pathways directly or indirectly associated with synaptic degeneration, G-protein coupled receptors (GPCR) signaling and immune responses. Interestingly, several distinct pathways associated with upregulated DEGs in VaD can be identified, including PTEN signaling, AMPK signaling, hepatic fibrosis and DNA damage response associated with BRCA1 (**Supplementary Table S5**).

### 3.6 DEGs indicating vascular dysfunction and neurodegeneration present in AD, VaD and MIX

Lastly, we identified Vascular Dysfunction and Neurodegeneration as the two main enrichment characteristics in AD, VaD and MIX using a combination of all DEGs to determine significant involvement of disease processes and biofunctions (**Figure 4C** and **Supplementary Table S6**). Vascular Dysfunction, as indicated by processes including “cerebrovascular dysfunction”, “hypertension”, “stroke” and “inhibition of development of vasculature”, was generally detected in all dementia groups. Interestingly, “arteriosclerosis”, “infarction”, as well as “inhibition of binding of vascular endothelial cells” were only detected in VaD, but not AD and MIX, consistent with a small vessel disease background of our VaD group. Furthermore, all dementia groups also detected Neurodegeneration, as indicated by “inhibition of neurotransmission”, inactivation of “neuritogenesis”, “dendritic growth/branching”, “long-term potentiation”, “excitatory postsynaptic potential”, “cognition”, “memory”, “synaptic depression”, as well as associations with “cerebral disorder”, “cognitive impairment”, “dementia”, “anxiety”, “emotional behaviours” and “AD” (**Figure 4C**).

## 4 DISCUSSION

In this head-to-head transcriptomics study of well-characterized Alzheimer’s disease, vascular dementia and mixed dementia neocortical tissues, we report on a wide range of shared and unique gene expression changes which both corroborate and extend previous work [14–19], and summarize below the wider themes and new insights gleaned from the data.

Firstly, the finding of Neurovascular Dysfunction and Neurodegeneration as the two main common processes in all three dementia groups (AD, VaD and MIX, see **Figure 4C** and **Supplementary Table S6**) is supported by an increasing recognition of the extensive interplay between vascular and neurodegenerative processes in dementia. For example, whilst neurodegenerative processes are well established drivers of dementia in AD and MIX, there is accumulating evidence that CeVDs, especially small vessel diseases, also cause neurodegeneration via multiple mechanisms including blood-brain barrier leakage, neurovascular unit dysfunction, hypoperfusion / hypoxia and neuroinflammation [50–52]. Indeed, cerebrovascular dysfunction is known to directly contribute to the onset and progression of AD [53, 54]. More recent findings using vasculature tissues for single-nuclei transcriptome sequencing demonstrated vascular cell type-specific perturbations in AD, as well as GWAS detection of at least 30 of the top 45 AD genes enriched in cells of human brain vasculature, suggest a pivotal role of cerebrovascular dysfunction in AD [55].

Of the pathways found to be affected in our cohort, the enrichment of extracellular matrix (ECM) organization and integrin signaling (**Supplementary Table S4**) were of particular interest, as activation of ECM and integrin-mediated adhesion and signaling may be associated with responses to vascular dysfunction in view of their crucial roles in neurovascular development and integrity [37]. We speculate that overactivation and excessive accumulation of ECM in cerebral vessels may affect intrinsic properties of the vessel wall (e.g., thickness and stiffness) leading to compromised cerebral blood flow and consequent impacts on cognitive function. Indeed, alterations in the cerebral vascular basement membrane have been reported in AD including thickening, altered composition and Aβ deposition [56].

Rho GTPase signaling is another pathway associated with cell migration, adhesion, cell shape, and proliferation, and is known to contribute to endothelial vasculogenesis, angiogenesis and mature vessel barrier function [40]. Interestingly, enrichment of Rho GTPase signaling from upregulated DEGs was detected in all dementia groups, whereas the enrichment from downregulated DEG was only detected in VaD but not in AD or MIX (**Supplementary Table S4**). We note that although Rho GTPase signaling remained significantly altered in all groups when using all DEGs for canonical pathway enrichment analysis, only VaD manifested inhibitory status as shown by negative Z-scores (**Supplementary Table S5**). Furthermore, IPA diseases and biofunctions enrichment analysis showed that impairment of development of vasculature, arteriosclerosis and infarction were only detected in VaD, but not in AD and MIX (**Supplementary Table S6**). Taken together, our data suggest that the underlying mechanisms of vascular dysfunction in VaD may differ from AD and MIX.

In this study, MIX demonstrated a substantially greater degree of overlap in DEGs with AD than with VaD (**Figure 1A**), even though MIX fulfilled diagnostic criteria for both AD and VaD. This is consistent with previous findings showing that the impacts of AD pathology outweighed vascular abnormalities in terms of influence on the severity and profile of cognitive impairments [12, 57, 58]. Additionally, higher proportions of downregulated DEGs and impaired canonical pathways, including mitochondrial dysfunction, were also detected in MIX compared with AD and VaD (**Figure 1A** and **Figure 4A**), suggesting that additive or synergistic deleterious effects from AD-related pathologies and vascular abnormalities may contribute to these outcomes in MIX, again consistent with previous findings [8, 58–60].

### 4.1 Study limitations

A usual requirement for comparative studies of dementia types is for disease and demographic variables, including severity of cognitive impairment, to be matched as much as possible across all dementia groups. In our cohort, VaD participants were of the subcortical ischemic subtype, which has been reported to manifest impairments of executive/ attention processing, contrasting with AD having more impairments in episodic memory [61]. Hence, MMSE was significantly higher in VaD as compared to AD (**Table 1**), likely due to the nature of the examination being weighted heavily towards episodic memory rather than executive function. Since there are various VaD subtypes, conclusions of this study are necessarily limited to subcortical ischemic subtype of VaD and MIX. Furthermore, to facilitate between-group comparisons, we set an arbitrary fold change cut-off of 1.5 with FDR<0.1 to determine DEG in each group, noting that this arbitrary cut-off, while in line with previous studies, may potentially lead to miscategorization of DEGs. To address this limitation, we focused more on overall pathway alterations with FDR correction rather than discussions based on specific DEGs. Finally, due to limited availability of samples and resources, we were not able to perform qPCR confirmation for all corresponding DEGs in altered pathways.

## 5 CONCLUSION

In this postmortem cortical transcriptomics study, we identified vascular dysfunction and neurodegeneration as the two major common processes in AD, VaD and MIX. Although VaD manifested the fewest DEGs, specific altered pathways associated with vascular dysfunction were identified. Additionally, whilst MIX showed substantial overlaps with AD, it was also enriched for uniquely altered DEGs suggestive of additive or synergistic deleterious effects arising from interactions of AD and vascular pathologies. Taken together, our study adds to current knowledge on shared and unique gene expression changes in the AD, VaD and mixed dementia cortex.

## Data Availability

The human transcriptome dataset has been deposited in Gene Expression Omnibus (GEO) data repository (Access #GSE164390). Other data that support the findings of this study are available on reasonable request from the corresponding authors.

https://www.ncbi.nlm.nih.gov/geo/query/acc.cgi?acc=GSE164390

## Abbreviations

Aβ42: β-amyloid_1-42_ peptide
AD: Alzheimer’s disease
ANOVA: analysis of variance
BA: Brodmann area
BRCA1: breast cancer gene 1
CTRL: non-demented aged controls
DEG: significant and differentially expressed genes
ECM: extracellular matrix
FDR: false discovery rate
GFAP: glial fibrillary acidic protein
GO: gene ontology
GWAS: genome-wide association study
HIF3A: hypoxia-inducible factor 3 alpha
IPA: Ingenuity Pathway Analysis
ITPKB: inositol-trisphosphate 3-kinase B
MIX: mixed AD and VaD
KEGG: Kyoto Encyclopedia of Genes and Genomes
MMSE: Mini-Mental State Examination
NFT: neurofibrillary tangles
NVU: neurovascular unit
PMI: Post-mortem interval
PTEN: phosphatase and tensin homolog
RT-qPCR: reverse transcription quantitative polymerase chain reaction
RIN: RNA Integrity Number
VaD: vascular dementia.

## CRediT Contributor Statement

Conceptualization: MGKT, MKPL; Data Curation: PTF, CB, RNK, MKPL; Investigation and Formal Analysis: FTWL, YLC, JHL, CYBL, MGKT; Funding Acquisition: CPC, MKPL; Resources: BKK, CPC, TML, MKPL, MGKT; Supervision: CPC, MKPL, MGKT; Visualization: FTWL, YLC, CYBL; Writing – Original Draft: MGKT, MKPL; Writing – Review & Editing: FTWL, YLC, JHL, CYBL, PTF, CB, RNK, BKK, CPC, TML, MKPL, MGKT

## Ethics approval and consent to participate

For the postmortem study, ethics approval for the collection and study of brain tissues received Institutional Review Board approval in both the UK (08/H1010/4) and Singapore institutions (NUS 12-062E), and informed consent was obtained from participants’ next-of-kin prior to removal of brain.

## Consent for publication

All authors gave consent for publication.

