## Supplementary Figure S1 for "Comparative cortical transcriptomic profiling of Alzheimer’s disease, vascular dementia and mixed dementia"

### Supplementary Figure S1. Correlations of selected transcriptomics-derived DEGs with RT-qPCR

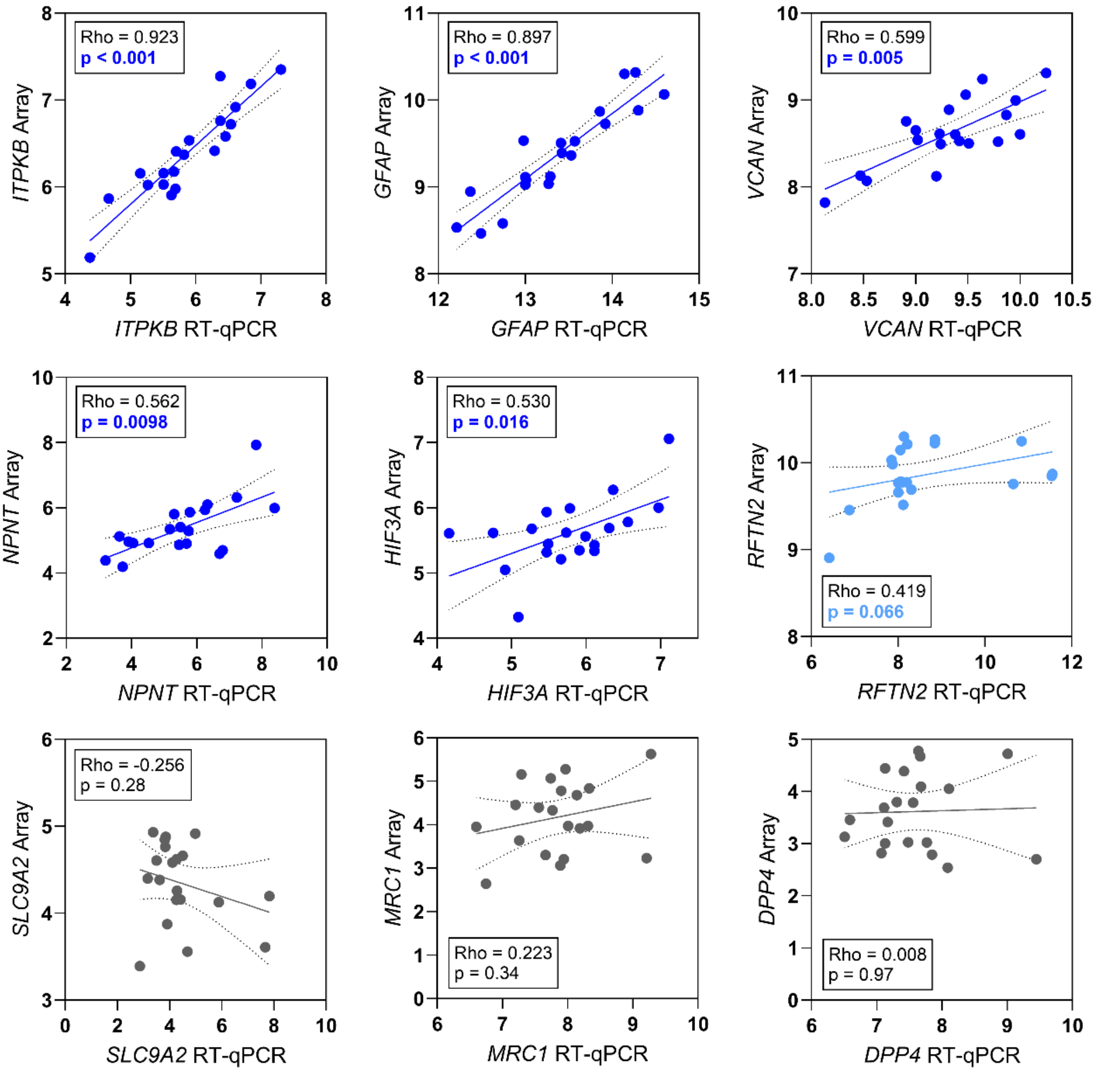


**Fig. S1.** Spearman correlations of selected genechip array-derived gene expression (Y-axes) with corresponding reverse transcription quantitative polymerase chain reaction (RT-qPCR) values (X-axes), both in arbitrary units. Insets denote rho coefficients and p-values. Data consisted of n=20 participants (3 CTRL, 5 VaD, 7 AD and 5 MIX). Genes measured are: *ITPKB,* inositol-trisphosphate 3-kinase B; *GFAP,* glial fibrillary acidic protein; *VCAN,* versican; *NPNT,* nephronectin; *HIF3A,* hypoxia inducible factor 3 subunit alpha; *RFTN2,* raftlin family member 2; *SLC9A2,* solute carrier family 9 member A2; *MRC1,* mannose receptor C-type 1; *DPP4,* dipeptidyl peptidase 4.
